# Derivation and validation of a non-invasive optoacoustic imaging biomarker for patients with intermittent claudication

**DOI:** 10.1101/2023.10.19.23297246

**Authors:** Milenko Caranovic, Julius Kempf, Yi Li, Adrian P. Regensburger, Josefine S. Günther, Anna P. Träger, Werner Lang, Alexander Meyer, Alexandra L. Wagner, Joachim Woelfle, Roman Raming, Lars-Philip Paulus, Adrian Buehler, Wolfgang Uter, Michael Uder, Christian-Alexander Behrendt, Markus F. Neurath, Maximilian J. Waldner, Ferdinand Knieling, Ulrich Rother

**Affiliations:** Department of Vascular Surgery, University Hospital Erlangen, Friedrich-Alexander-Universität Erlangen-Nürnberg (FAU), Krankenhausstraße 12, D-91054 Erlangen, Germany; Department of Pediatrics and Adolescent Medicine, University Hospital Erlangen, Friedrich-Alexander-Universität Erlangen-Nürnberg (FAU), Loschgestraße 15, D-91054 Erlangen, Germany; Department of Vascular and Endovascular Surgery, University Hospital Münster, Westfälische Wilhelm-Universität Münster (WWU), Albert-Schweitzer-Campus 1, D-48149 Münster, Germany; Department of Medical Informatics, Biometry and Epidemiology, Friedrich-Alexander-Universität Erlangen-Nürnberg (FAU), Waldstraße 6, D-91054 Erlangen, Germany; Institute of Radiology, University Hospital Erlangen, Friedrich-Alexander-Universität Erlangen-Nürnberg (FAU), Maximiliansplatz 1, D-91054 Erlangen, Germany; Department of Vascular and Endovascular Surgery, Asklepios Klinik Wandsbek, Asklepios Medical School, Alphonsstraße 14, D-22043 Hamburg, Deutschland; Department of Medicine 1, University Hospital Erlangen, Friedrich-Alexander-Universität Erlangen-Nürnberg (FAU), Ulmenweg 18, D-91054 Erlangen, Germany; Faculty of Medicine, Friedrich-Alexander-Universität Erlangen-Nürnberg (FAU), Krankenhausstraße 12, D-91054 Erlangen, Germany; Deutsches Zentrum für Immuntherapie (DZI), University Hospital Erlangen, Friedrich-Alexander-Universität Erlangen-Nürnberg (FAU), Ulmenweg 18, D-91054 Erlangen, Germany; Erlangen Graduate School in Advanced Optical Technologies (SAOT), Friedrich-Alexander-Universität Erlangen-Nürnberg, Paul-Gordan-Straße 6, D-91052 Erlangen, Germany

**Keywords:** Peripheral artery disease, Intermittent Claudication, Microcirculation, MSOT, Optoacoustic imaging

## Abstract

**Background:** Multispectral optoacoustic tomography (MSOT), a molecular sensitive ultrasound, offers a non-invasive diagnostic approach to image the deep-tissue biomarkers.

**Objectives:** The authors aimed to investigate the diagnostic accuracy of MSOT to distinguish between healthy volunteers (HV) and patients with intermittent claudication (IC) by assessing hemoglobin-related biomarkers in calf muscle tissue.

**Methods:** In this monocentric, cross-sectional diagnostic trial using derivation (DC) and validation cohorts (VC) yll subjects underwent standardized PAD diagnostics. This included pulse palpation, ankle brachial index (ABI), duplex sonography, 6-minute walk test (6MWT), and assessment of health-related quality of life (VASCUQOL-6). The vascular occlusion profile in IC patients was confirmed by angiography (aggregated TransAtlantic Inter-Society Consensus classification, aTASC). MSOT imaging of calf muscle was performed before and after standardized heel raise provocation.

**Participants:** Of 123 screened individuals, 102 completed the study. MSOT-derived oxygenation (msO_2_) after the exercise differentiated IC and HV with an area under curve the receiver operator characteristics curve (AUROC) in DC by 0.99 (95%CI 0.97;1.00, p<0.001, sensitivity: 100%, specificity: 95.8%) and in the VC by 0.95 (95%CI 0.95;1.00, p<0.001, sensitivity: 96.2%, specificity: 96.0%). mSO_2_ positively correlated with the ABI post-exercise (R=0.83, 95%CI 0.75;0.88, P<0.001), the absolute walking distance in the 6MWT (R=0.77, 95%CI 0.68;0.84, P<0.001), the VASCUQOL-6 (R=0.79, 95%CI 0.70;0.85, P<0.001) and negatively with aTASC classification (R=-0.80, 95%CI -0.86;-0.72, P<0.001).

**Conclusions:** Post-exercise MSOT-derived saturation in the calf muscle was validated as a new and promising diagnostic biomarker to distinguish between HV and IC patients yielding high sensitivity and specificity. (<u>NCT05373927</u>)

## Introduction

With 237 million cases worldwide, peripheral artery disease (PAD) ranges among the three most common cardiovascular diseases (1). The occlusion of vessels, as seen in PAD, can progress asymptomatically and finally result in symptoms like intermittent claudication (IC), rest pain or ischemic ulceration (2,3). First-line diagnostics includes palpation of pulses, measurement of the blood pressure derived ankle-brachial index (ABI), or color-coded duplex sonography. Other non-invasive diagnostic modalities are mainly used in later stages, which are for example transcutaneous oxygen pressure (tcPO2), laser-doppler flowmetry, skin-perfusion pressure of fluorescence angiography (2,3). However, to confirm the ischemic origin of symptoms, invasive angiographic imaging procedures are often required.

Despite the existence of these various diagnostic options, patients with unclear leg pain during walking remain challenging for diagnosis (4). In these cases, a diagnostic tool for direct assessment of the affected organ, namely the lower leg musculature in PAD, would be helpful.

In this regard, Multispectral Optoacoustic Tomography (MSOT) enables visualization and quantification of muscle tissue properties, such as hemoglobin of different oxygenation states over a broad range of applications (5–8). MSOT has already demonstrated feasible to detect differences in terms of hemoglobin saturation of the calf muscle between healthy volunteers (HV) and different stages of PAD (9). However, its diagnostic accuracy to differentiate between healthy and early claudication stage, which would be clinically relevant, is still elusive.

Therefore, this study aimed to develop appropriate thresholds and subsequently validate the diagnostic performance of MSOT to discriminate HV from IC patients by assessing the calf muscle before and after exercise.

## Patients and Methods

### Trial Overview

The prospective data collection and analysis was designed as a monocentric, cross-sectional diagnostic study and performed at the Department of Vascular Surgery of the University Hospital Erlangen, Germany, between 9^th^ May 2022 and 14^th^ July 2022. The trial was approved by the local ethics committee of the University Erlangen-Nürnberg (22-62-Bm) and registered on clinicaltrials.gov (NCT05373927).

A total of 52 healthy volunteers (HV) and 50 IC patients were recruited. In the derivation cohort (DC), 27 HV were compared to 24 IC patients. HV were required to be at least 50 years old, have palpable foot pulses and a normal ABI (values between 0.9 and 1.4). Exclusion criteria were existing diabetes mellitus, chronic renal insufficiency, and IC symptoms. IC patients were included in Fontaine stage IIa/IIb or Rutherford category 1 to 3. The findings in the DC were applied to an independent validation cohort (VC, 25 HV vs 26 IC) to confirm the findings.

### Trial Procedures

Potential participants were screened for inclusion and exclusion criteria and if suitable for the study, they were informed about the procedure, benefits, and risks. After giving written consent, examinations were done (exactly comparable between HV and IC, with except of angiography) during one single appointment within 90 minutes. Anamnesis was taken, in which all essential information related to PAD diagnosis was obtained (see Supplementary Appendix). The standardized VASCUQOL-6 questionnaire was used to assess the individual PAD-specific quality of life (see Supplementary Appendix). A standardized non-invasive routine examination for PAD diagnostics was performed, which included pulse palpation, ABI before and after exercise and color-coded duplex ultrasound (CCDS)(3). The vascular occlusion profile in IC patients was confirmed by angiographic imaging (either magnetic resonance angiography, computed tomography angiography or digital subtraction angiography). All MSOT measurements were performed with the same CE-certified system (Acuity Echo, iThera Medical GmbH, Munich) at wavelengths between 700 and 1210 nm. By using the integrated B-mode ultrasound function, the optimal position for imaging the triceps surae muscle was determined in the B-mode scan. The final position of the transducer was marked and the exact same position was retrieved for the pre- and post-exercise measurement. The MSOT parameters deoxygenated hemoglobin (Hb), oxygenated hemoglobin (HbO_2_) and MSOT saturation (mSO_2_) were then determined for the muscle (exemplary MSOT images can be found in figure 3). The first measurement was taken after a rest of 10 minutes in a lying position. After this initial measurement, all participants performed the heel raise exercise to strain the calf muscle, repeated until pain occurred. Directly after the exercise, another ABI measurement and the second MSOT measurement at the marked position of the calf muscle were performed.

After a resting phase of at least 30 minutes, the participants underwent a Six-Minute Walk Test (6MWT) to determine the relative (distance until the first pain occurs), absolute (distance until the first stop due to calf pain) and total (distance walked in six minutes) walking distance.

### Data Analysis

The examiners for the final MSOT data analysis were blinded to the results of the clinical assessments. All data processing for measurements of imaging parameters before and after the heel raise exercise was performed by iLabs software (Version 1.2.9, iThera Medical GmbH, Munich). As demonstrated before, deoxygenated hemoglobin (Hb), oxygenated hemoglobin (HbO2) and MSOT-derived oxygenation (mSO_2_) were assessed after drawing a standardized region of interest (ROI) directly below the muscle fascia of the triceps surae muscle (9). The Hb and HbO_2_ values were obtained directly from the generated data using an algorithm, while msO_2_ equals the quotient of HbO_2_ by Hb+HbO_2_. It should be emphasized that the parameter described as msO_2_ does not correspond to the actual physical oxygen saturation and should therefore be considered as an independently generated parameter. MSOT values are given in arbitrary units (a.u.). Angiographic images of the IC patients were reviewed by two independent investigators (M.C., J.K.) and categorized according to the trans-atlantic inter-society consensus document II (TASC)(10,11). In the event of uncertainties, a third opinion (U.R.) was consulted for a collaborative decision-making. The classification evaluates the anatomic distribution and extent of lower extremity vascular lesions at three separate levels: Aorto-iliac, femoral popliteal and infrapopliteal. In order to obtain an overview of the functional severity of the PAD, we used an additional classification (aTASC, see Supplementary Appendix), which combines the separately reviewed areas and considers the collateralization situation. The additional classification includes three classes (aTASC1: angiographically healthy, aTASC 2: good collateralization capability, aTASC 3 poor collateralization capability).

### Statistical analyses

Based on data collected of a previous study cohort, we estimated a minimum sample size of 11 (oxygenated hemoglobin [HbO_2_])-33 (deoxygenated hemoglobin [Hb]) participants in each clinical group (HV, IC) (80% power, alpha of 0.05 one-sided). Continuous variables are given as mean values with standard deviation. Categorical factors are presented as absolute and relative frequencies. Correlations are given by Pearson’s correlation coefficient (R). Single missing values led to exclusion of subjects for the specific analysis. In case no muscle was appropriately imaged in the MSOT frames, the subject was excluded from all further analyses. For all statistical tests p<0.05 was considered statistically significant. Analyses were performed using IBM SPSS Statistics (version 28, IBM Corp., N.Y., USA), R Statistics (version 4.0.5.) and GraphPad PRISM (version 9.4.1, GraphPad Software, USA). Statistical differences of DC and VC were tested by Fisher test or Welch test. To illustrate the diagnostic quality to differentiate between HV and IC by MSOT, the sensitivity, specificity, area under the curve (AUC) and the cut-off point (derived from the optimal Youden index) were determined in the DC. The cut-off point was then applied to the VC to validate the results from the DC. Estimates were accompanied by 95% confidence limits (95% CIs).

## Results

### Patient characteristics

A total of 123 individuals were consecutively screened for enrollment between 9^th^ May 2022 and 14^th^ July 2022, of whom 115 gave written informed consent. In 109 cases, the study was completed with a full data set and finally 102 of them were used for data processing (compare study flow chart, **Figure 1**). HV had a mean age 63.8 ± 8.0 years and 45.1% were female. Patients with IC were 60.0 ± 6.0 years of age and 42.3% were female. A description of the patient characteristics is given in **Table 1**. We found no significant difference between the characteristics of the consecutively collected separated DC and VC (see Supplementary Appendix).

**Figure 1.**
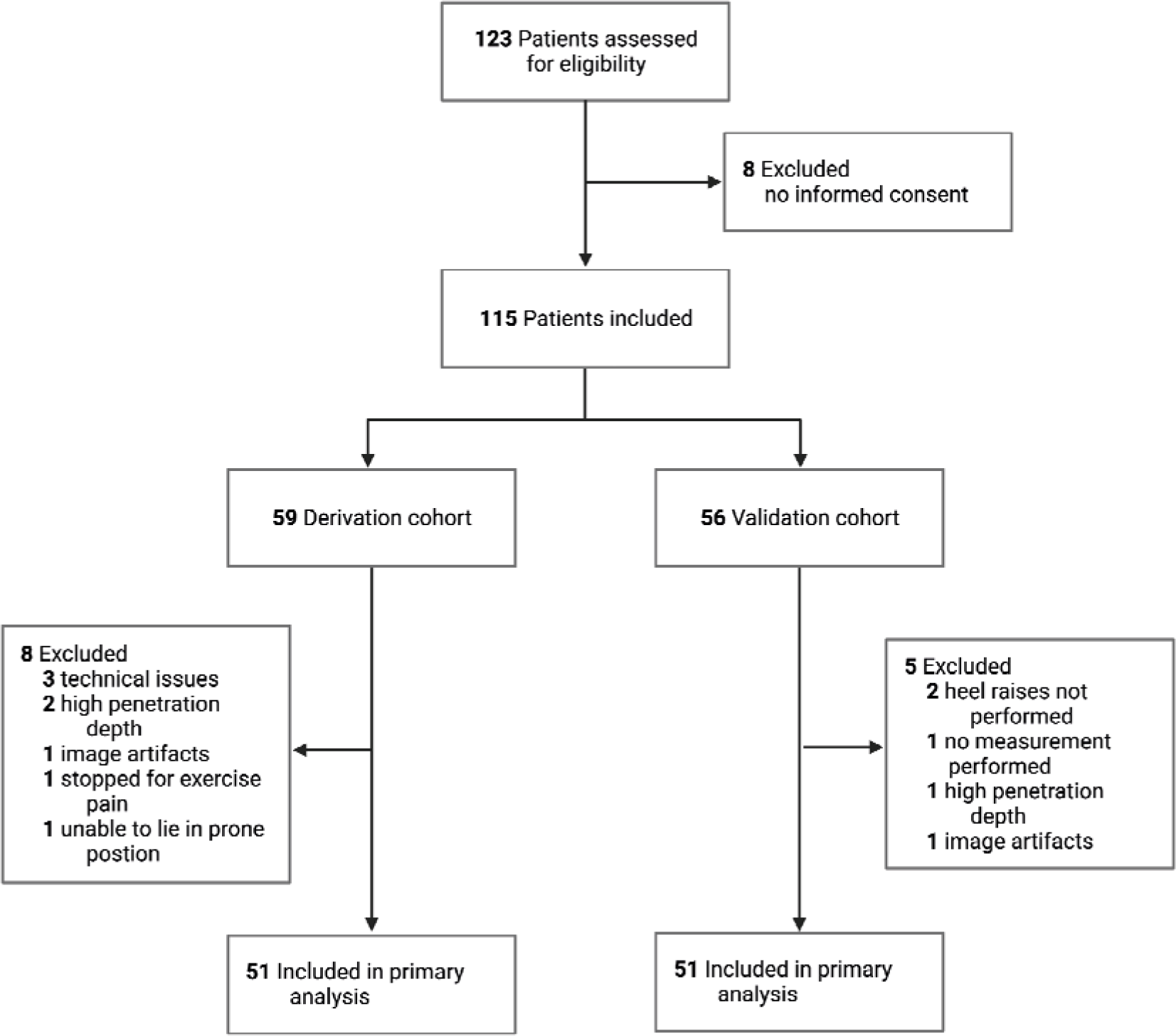
Flow chart of patient recruitment and data collection process.

**Table 1.**
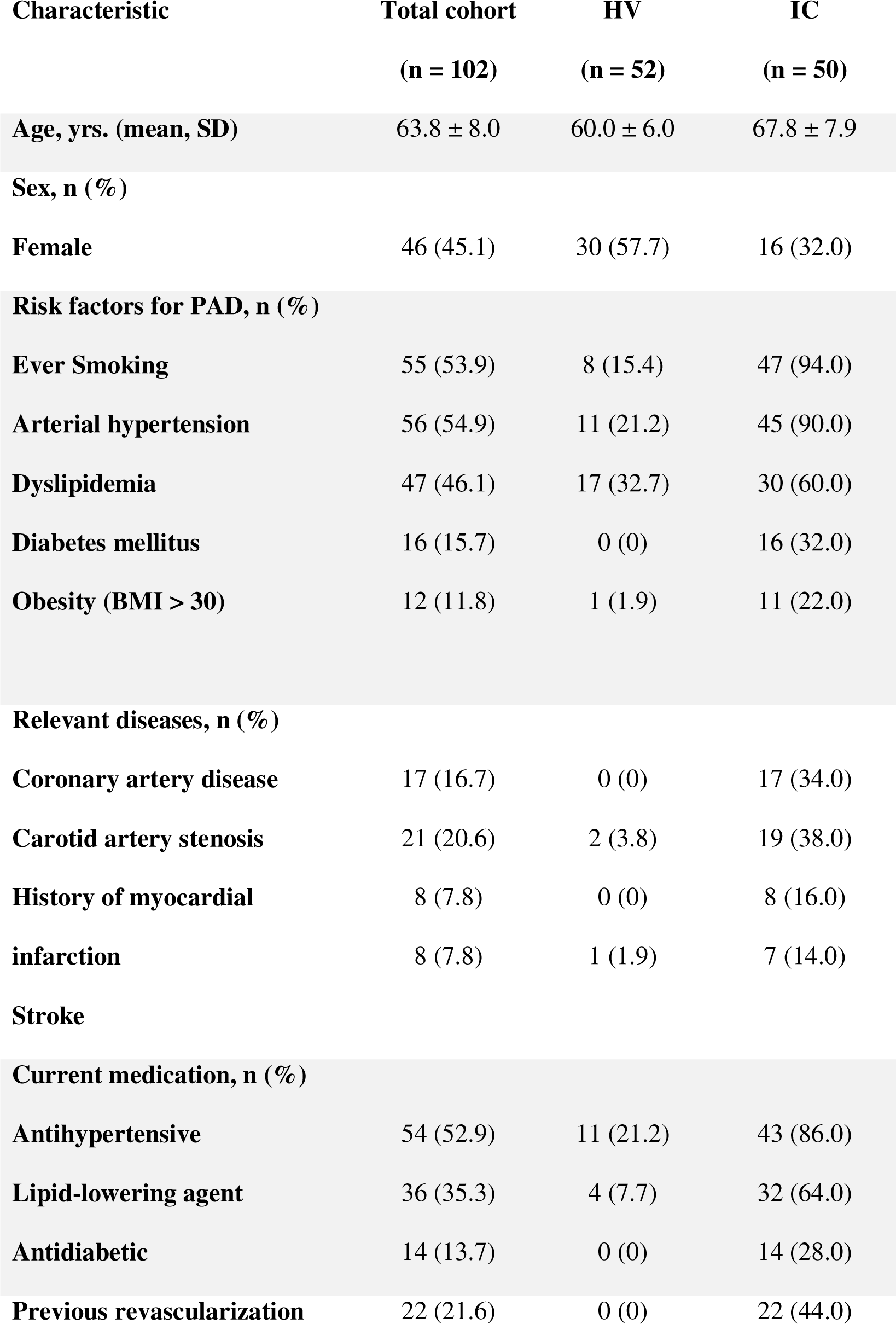
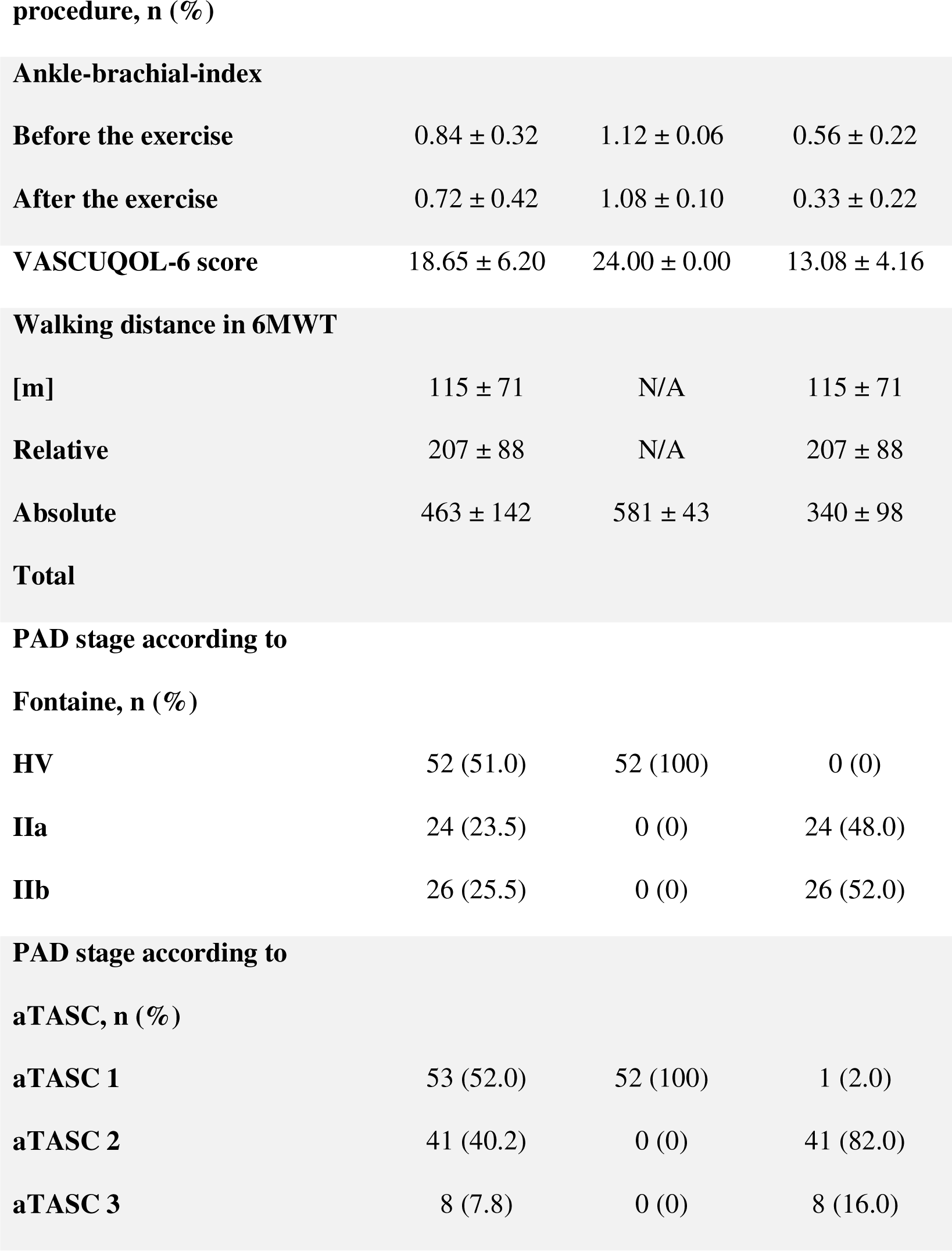
Demographic and Clinical Characteristics. Categorical data represented by absolute and relative frequencies, continuous data by mean ± standard derivation. Statistical comparison of the HV and IC and total cohort using the Chi-squared test for categorical data and the t-test for continuous data. (HV, healthy volunteers; IC, patients with peripheral arterial disease with in Fontaine stage IIa/IIb or Rutherford category 1 to 3; VASCUQOL-6, Vascular Quality of Life Questionnaire-6; 6MWT, Six-Minute-Walk-Test; PAD, peripheral arterial disease; aTASC (Type 1: HV or IC with no signs of stenosis or occlusion in angiography; Type 2: signs of stenosis or occlusion in the femoropopliteal and/or infrapopliteal area or TASC-II-level A or B in the aortoiliac area; Type 3: TASC-II-level C or D in the aortoiliac area); N/A, not applicable)

| Characteristic | Total cohort<br>(n = 102) | HV<br>(n = 52) | IC<br>(n = 50) |
| --- | --- | --- | --- |
| Age, yrs. (mean, SD) | 63.8 ± 8.0 | 60.0 ± 6.0 | 67.8 ± 7.9 |
| Sex, n (%) |  |  |  |
| Female | 46 (45.1) | 30 (57.7) | 16 (32.0) |
| Risk factors for PAD, n (%) |  |  |  |
| Ever Smoking | 55 (53.9) | 8 (15.4) | 47 (94.0) |
| Arterial hypertension | 56 (54.9) | 11 (21.2) | 45 (90.0) |
| Dyslipidemia | 47 (46.1) | 17 (32.7) | 30 (60.0) |
| Diabetes mellitus | 16 (15.7) | 0 (0) | 16 (32.0) |
| Obesity (BMI > 30) | 12 (11.8) | 1 (1.9) | 11 (22.0) |
| Relevant diseases, n (%) |  |  |  |
| Coronary artery disease | 17 (16.7) | 0 (0) | 17 (34.0) |
| Carotid artery stenosis | 21 (20.6) | 2 (3.8) | 19 (38.0) |
| History of myocardial infarction | 8 (7.8) | 0 (0) | 8 (16.0) |
| Stroke | 8 (7.8) | 1 (1.9) | 7 (14.0) |
| Current medication, n (%) |  |  |  |
| Antihypertensive | 54 (52.9) | 11 (21.2) | 43 (86.0) |
| Lipid-lowering agent | 36 (35.3) | 4 (7.7) | 32 (64.0) |
| Antidiabetic | 14 (13.7) | 0 (0) | 14 (28.0) |
| Previous revascularization | 22 (21.6) | 0 (0) | 22 (44.0) |
| **procedure, n (%)** |  |  |  |
| **Ankle-brachial-index** |  |  |  |
| **Before the exercise** | 0.84 ± 0.32 | 1.12 ± 0.06 | 0.56 ± 0.22 |
| **After the exercise** | 0.72 ± 0.42 | 1.08 ± 0.10 | 0.33 ± 0.22 |
| **VASCUQOL-6 score** | 18.65 ± 6.20 | 24.00 ± 0.00 | 13.08 ± 4.16 |
| **Walking distance in 6MWT** |  |  |  |
| **[m]** | 115 ± 71 | N/A | 115 ± 71 |
| **Relative** | 207 ± 88 | N/A | 207 ± 88 |
| **Absolute** | 463 ± 142 | 581 ± 43 | 340 ± 98 |
| **Total** |  |  |  |
| **Fontaine, n (%)** |  |  |  |
| **HV** | 52 (51.0) | 52 (100) | 0 (0) |
| **IIa** | 24 (23.5) | 0 (0) | 24 (48.0) |
| **IIb** | 26 (25.5) | 0 (0) | 26 (52.0) |

**Table 1 - Demographic and Clinical Characteristics**
|  |  |  |  |
| --- | --- | --- | --- |
| **aTASC 1** | 53 (52.0) | 52 (100) | 1 (2.0) |
| **aTASC 2** | 41 (40.2) | 0 (0) | 41 (82.0) |
| **aTASC 3** | 8 (7.8) | 0 (0) | 8 (16.0) |

### Diametral response of MSOT-derived parameters in HV and IC

All patients underwent standardized MSOT imaging before and after exercise (**Figure 2A**). At resting state in the DC, we found no significant difference between the HV and IC group for the parameters Hb (0.15±0.05a.u. vs. 0.18±0.05a.u; P=0.79) and HbO_2_ (0.19±0.06a.u. vs. 0.18± 0.05a.u.; P=0.42). A significant difference was observed for mSO_2_ (0.56± 0.04a.u vs. 0.50 ± 0.04 a.u.; P<0.001). After heel raise exercise, Hb increased in IC patients (0.17±0.06a.u. vs. 0.27± 0.06a.u.; P<0.001). Opposite to Hb, HbO_2_ increased in the HV (0.26±0.06a.u. vs 0.182±0.03a.u.; P<0.001). Given this disease-dependent diametric response, mSO_2_ logically also differentiated IC and HV group (0.41±0.06a.u. vs. 0.61 ± 0.04a.u.; P< 0.001) (**Figure 2B**). Exemplary MSOT images are given in **Figure 2C**.

**Figure 2.**
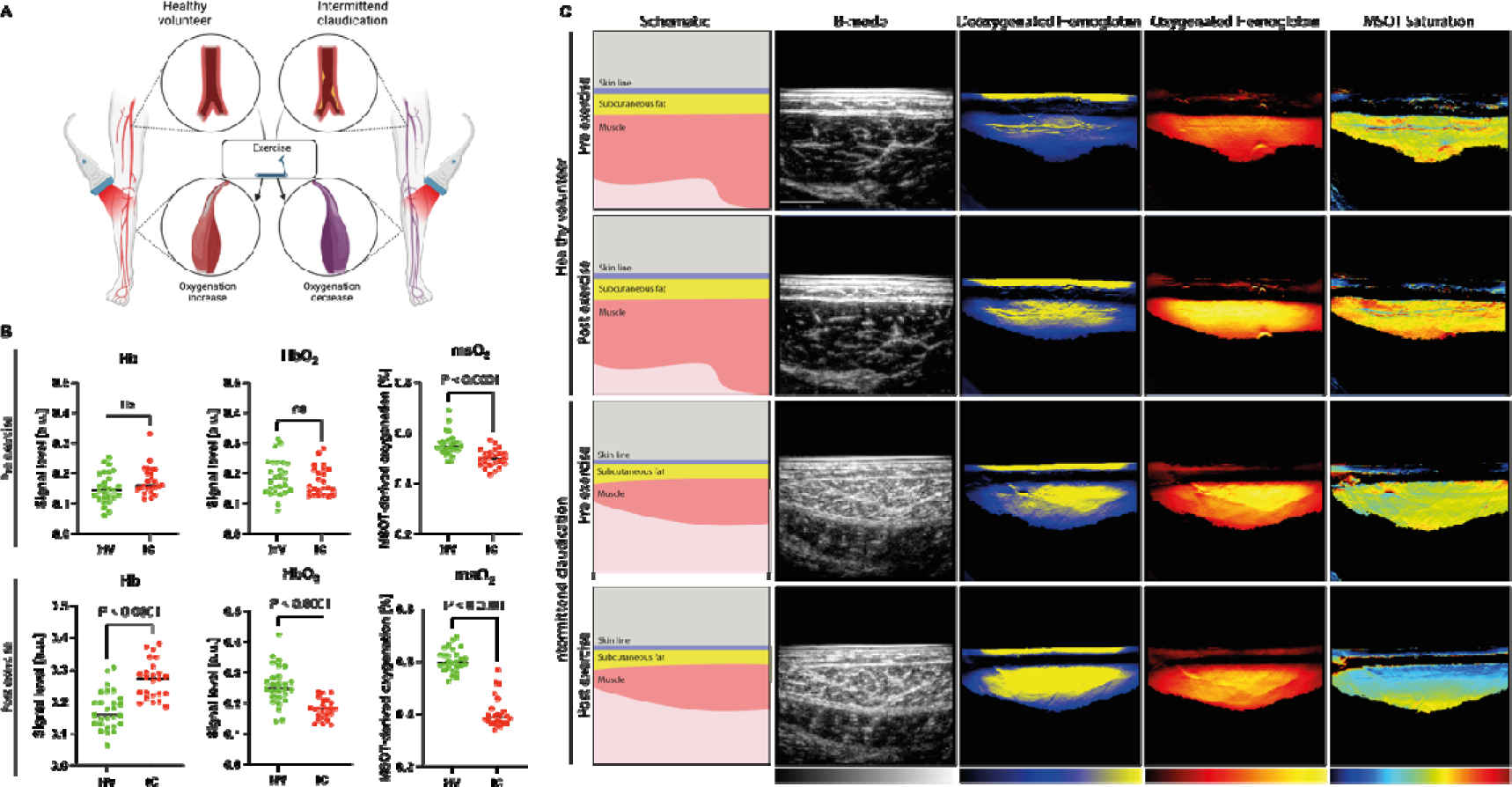
MSOT images of a HV and an IC before and after the heel raise exercise. MSOT Imaging approach in the study (**A**). Dot plots of all patients in the derivation cohort pre- and post-exercise (**B**). MSOT images of a healthy volunteer (HV) and a patient with peripheral arterial disease in Fontaine stage IIa/IIb or Rutherford category 1 to 3 (IC) before (Pre) and after (Post) the heel raise exercise showing the parameters deoxygenated hemoglobin (Hb), oxygenated hemoglobin (HbO_2_) and MSOT saturation (mSO_2_). The first column shows a schematic representation of the visible anatomical structures (skin line, subcutaneous fat, muscle); the second shows the sonographic image in B-mode. The following columns illustrate the MSOT data of Hb, HbO_2_ and msO_2_, visually represented as a heat map, with the colors reflecting the signal intensity (Hb: dark blue, minimum signal; yellow, maximum signal; HbO_2_: dark red, minimum signal; yellow, maximum signal; mSO_2_: dark blue, minimum signal; dark red, maximum signal) (**C**). Figure created with bioRender.com

### Diagnostic properties of MSOT to distinguish HV and IC

To assess the diagnostic properties of MSOT to distinguish HV and IC we calculated the receiver operating characteristics (ROC). At resting state, the corresponding area-under-the-curve (AUC) for Hb was 0.65 (95%CI 0.49;0.80), for HbO_2_ was 0.58 (95%CI 0.42;0.74), and was mSO_2_ 0.85 (95%CI 0.74;0.96) (**Figure 2A-C**). After heel raise exercise, the AUC of Hb increased to 0.87 (95%CI 0.77;0.97). The associated sensitivity in the DC was 74.1% (95%CI 55.3-86.8%) and the specificity 91.7% (95%CI 74.2-98.5%). Similarly, we found a sensitivity of 76.9% (95%CI 58.0-89.0%) and a specificity of 72.0% (95%CI 52.4-85.7%) in the VC (cut-off: 0.20a.u.) (**Figure 2D**). Opposite to Hb, were HbO_2_ increased in the HVs, the AUC was 0.87 (95%CI 0.76;0.97), with a sensitivity of 66.7% (95%CI 47.8-81.4%) and a specificity 100% (95%CI 86.2-100%) in the DC. This was validated with a sensitivity of 100% (95%CI 87.1-100%) and a specificity of 72.0% (95%CI 52.4-85.7%) (cut-off: 0.24a.u.) in the VC (**Figure 2E**). The quotient of the two previous parameters, mSO_2_ had an area under ROC curve of 0.99 (95%CI 0.97;1.00), with a sensitivity of 100% (95%CI 87.5-100%) and a specificity of 95.8% (95%CI 79.8-99.8%) (cut-off: 0.52a.u.). In the VC, a sensitivity of 96.2% (95%CI 81.1-99.8%) and a specificity of 96.0% (95%CI 80.5-99.8%) was confirmed (**Figure 2F**). A complete overview divided in DC and VC showing the described parameters before and after the exercise is given in the Supplementary Appendix (**Table S3**).

### Correlation to clinical standard assessments

Using the merged cohort (DC and VC, n=102, with except of aTASC, where only IC patients were assessed), the correlation between MSOT parameters (Hb, HbO_2_ and msO2) before and after the heel raise exercise were analyzed (Figure 3). At resting state, Hb, HbO_2_ and mSO_2_ showed low correlation to standard assessments (range of R ABI: -0.07 to 0.45, 6MWT: -0.11 to 0.45, VASCUQOL6: -0.09 to 042, aTASC: 0.06 to -0.49, all p>0.05).

**Figure 3.**
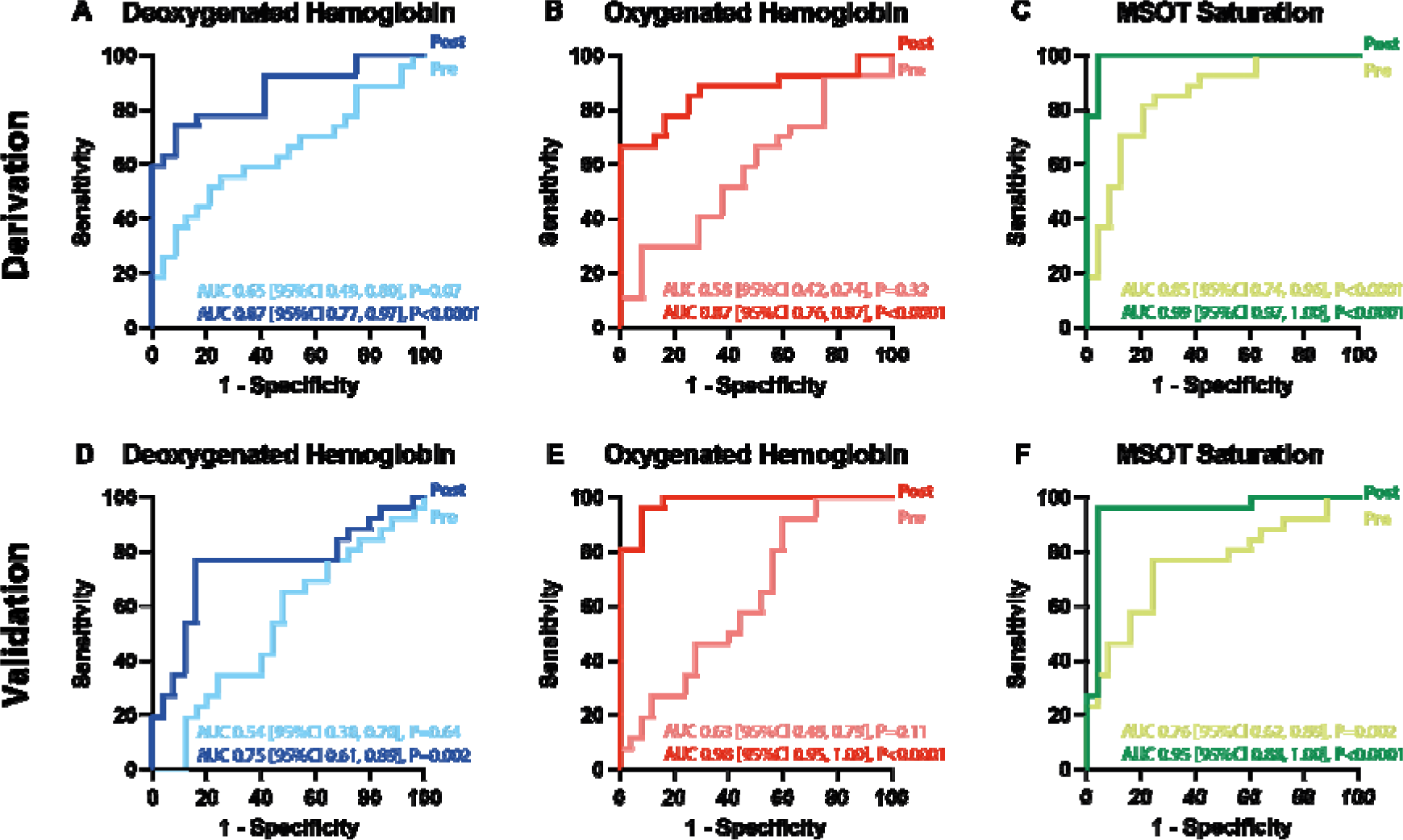
ROC analysis of the MSOT parameters Hb, HbO_2_ and msO_2_ in the DC. Receiver operating characteristic (ROC) analysis comparing the area under the curve (AUC) with 95% confidence interval (CI) of the MSOT parameters deoxygenated hemoglobin (**A**: Hb, blue curves), oxygenated hemoglobin (**B**: HbO_2_, red curves) and MSOT saturation (**C**: mSO_2_, green curves) of patients with peripheral arterial disease in Fontaine stage IIa/IIb or Rutherford category 1 to 3 (IC) and healthy volunteers (HV) before (Pre, light curve) and after (Post, dark curve) the heel raise exercise in the derivation cohort (DC). Hb (**D**), HbO2 (**E**), mSO_2_ (**F**) of the validation cohort (VC) pre-(light) and post-exercise (dark curve).

After exercise, Hb negatively correlated with ABI (R=-0.47, 95%CI -0.61;-0.30, P<0.001), 6MWT (R=-0.5095%CI -0.63;-0.33, P<0.001), VASCUQOL6 (R=-0.5195%CI -0.64;-0.35, P<0.001) and positively to aTASC (R=0.43, 95%CI 0.25;0.57, P<0.001). By contrast, HbO_2_ positively correlated with ABI (R=0.72, 95%CI 0.61;0.80, P<0.001), 6MWT (R= 0.61, 95%CI 0.47;0.72, P<0.001), VASCUQOL6 (R=0.63, 95%CI 0.49;0.73, P<0.001) and negatively to aTASC (R=-0.69, 95%CI -0.78;-0.57, P<0.001), the latter correlation analysis restricted to the IC subsample. Finally, mSO_2_ showed a strong positive correlation with the ABI after the exercise (R= 0.83, 95%CI 0.75;0.88, P<0.001), the absolute walking distance in the 6MWT (R=0.77, 95%CI 0.68;0.84, P<0.001), the VASCUQOL-6 questionnaire (R=0.79, 95%CI 0.70;0.85, P<0.001) and negative correlation with angiographic findings, stratified according to the aggregated TASC classification (R=-0.80, 95%CI -0.86;-0.72, P<0.001) (**Figure 4**).

**Figure 4.**
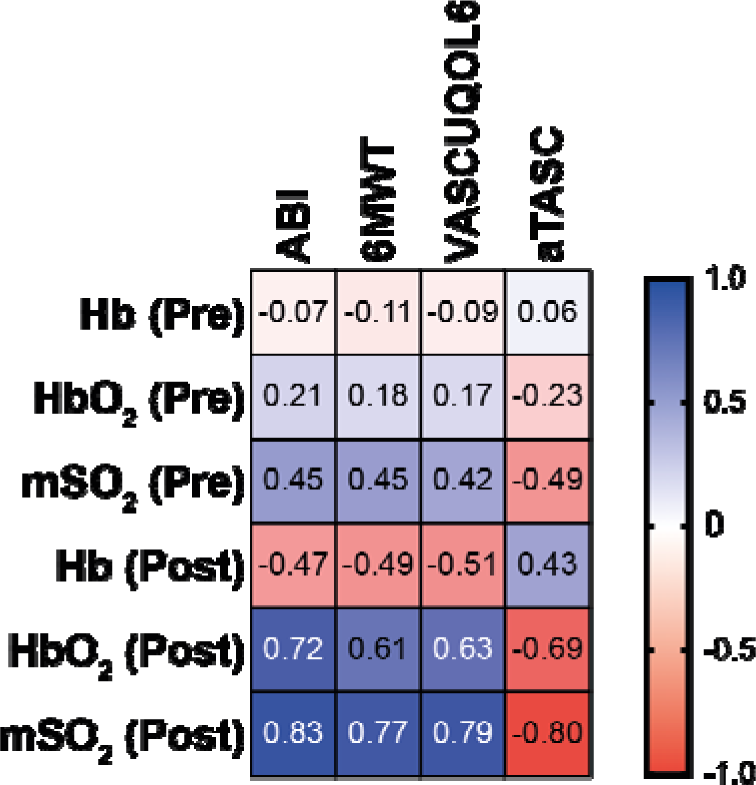
Correlation of MSOT parameters with clinical assessments. Correlation matrix of the MSOT parameters deoxygenated hemoglobin (Hb), oxygenated hemoglobin (HbO_2_), MSOT saturation (mSO_2_) before (Pre) and after (Post) heel raise exercise with ankle-brachial-index after heel raise exercise (ABI), the absolute walking distance in the Six-Minute Walk Test (6MWT), the score in the Vascular Quality of Life Questionnaire-6 (VASCUQOL-6), the score in the aggregated Trans-Atlantic Inter-Society Consensus Document on Management of Peripheral Arterial Disease classification II (aTASC). Correlation given by Pearson correlation coefficient (R) are indicated as strongly positive (dark blue) and strongly negative (dark red).

### Adverse events

No adverse events were observed.

## Discussion

In this study, three different hemoglobin parameters were derived with MSOT in the triceps surae muscle: Hb, HbO_2_ and mSO_2_. Using an easy to implement heel raise exercise, diverging MSOT signal responses in the HV and IC groups provided a strong differentiability between healthy volunteers and diseased patients. While diseased patients showed an increase in Hb and a decrease in HbO_2_, healthy volunteers showed the exact opposite reaction. This is why, the parameter mSO_2_, which is calculated through the quotient of HbO_2_ and Hb+HbO_2_, provided a strong differentiator for both groups. The data was generated in a derivation cohort and verified with high sensitivity and specificity in the independent validation cohort.

Since the photoacoustic effect has been described by Alexander Graham Bell in the late 19^th^ century (12), multiple developments transferred this technique to modern clinical diagnostic tools like MSOT (13). Similar technologies have already been used in clinical applications such as Crohn’s disease (14,15) or neuromuscular disorders (6,16). Furthermore, we already demonstrated its applicability for PAD diagnostic to grade different stages of the disease on based on different levels of calf muscle oxygenation (9). In this first initial trial, the diagnostic accuracy to discriminate between HV and IC patients was modest. The current significant increase of sensitivity and specificity could be achieved by examining responses to an easy-to-perform exercise-induced challenge prior to imaging.

Heel raise exercises have capability to lead to an exhaustion in the calf muscle even in healthy subjects, while this cannot commonly be reached by simple treadmill exercise. This challenge has proven its usability in PAD patients, as 30 seconds of heel raise exercise showed comparable post-exercise ABI results to 1 minute treadmill walking tests (4km/h, gradient of 10 degree) (17). A further advantage of the heel raise exercise is, that in a future perspective, this exercise can easily be implemented even in the outpatient setting to assess IC patients. MSOT muscle imaging is of certain interest, as it enables the direct assessment of endo-organ involvement in patients with intermittent claudication. The current diagnostic procedures in such diseases with various origins are mainly driven by treadmill or the 6MWT, assessing the pre- and post-exercise ABI (3,18). However, these tests do not objectify the degree of muscle impairment, and are often unable to give conclusive decision about the origin of the symptoms. Additionally, ABI is flawed with the known problems in diabetics and patients with chronic kidney disease, by which MSOT is not affected, as shown in the previous study (9).

Audonnet et al. used transcutaneous oxygenation measurements before and after treadmill exercise to distinguished claudication from lower back pain (19). However, the diagnostic potential of this method in this indication remains limited as it can merely investigate the skin oxygenation. This is, however, indirectly influenced by a competitive flow due to increase need of the muscle. MSOT has the capability to investigate the muscle directly in different locations of the body up to a depth of three centimeters, and might therefore help in cases of unclear diagnosis due to overlapping symptoms.

The clinical usability of the MSOT investigation might be predominantly in patients with IC. In this patient cohort, there are often patients with concomitant diseases, especially lumbar ischialgia, hip disorders or even in buttock claudication in which differential diagnosis can be challenging (4). In this context, the correlation of the MSOT derived values and the results of the 6MWT as well as the health-related quality of life, assessed by the VASCUQUOL 6, seem to be especially interesting, as these results underline the potential of the method in IC patients (20). Therefore, MSOT might be proposed as an adjunct measurement in patients with IC, as it can facilitate bedside diagnosis in distinct patients.

## Study limitations

This study has several limitations. Even though the study contains a derivation and validation cohort, it was conducted in a monocentric setting. Therefore, the results have to be confirmed in a multicenter setting to address reproducibility. In regards to technical limitations, deeper calf musculature can currently not be assessed in a reproduceable manner, which led to exclusion of 3 patients in total in this study.

## Conclusions

In conclusion, the MSOT-derived saturation in the post-exercise calf muscle was confirmed as a new diagnostic biomarker to distinguish between HV and IC patients with high sensitivity and specificity. This opens new diagnostic possibilities to ensure the ischemic origin of symptoms especially in diagnostic challenging patient cohorts due to various concomitant diseases.

## Data availability

The full study protocol and reporting summaries are available on the registry of ClinicalTrials.gov (ClinicalTrials.gov Identifier NCT05373927, Protocol Identifier MSOT_IC). The datasets generated and analyzed during the study are available from the corresponding authors on reasonable request. Restrictions may be valid due to patient privacy and the General Data Protection Regulation.

## Acknowledgements

The present work was performed in fulfillment of the requirements for obtaining the degree “Dr. med.”(M.C.).

## Abbreviations

ABI: Ankle-brachial-index
aTASC: Aggregated TASC II classification
a.u.: Arbitrary units
AUC: Area under the curve
CCDS: Color-coded duplex sonography DC Derivation Cohort
Hb: Deoxygenated hemoglobin
HbO_2_: Oxygenated hemoglobin
HV: Healthy volunteer(s)
IC: Intermittent claudication
MSOT: Multispectral Optoacoustic Tomography
mSO_2_: MSOT saturation
PAD: Peripheral artery disease
ROC: Receiver operating characteristic
ROI: Region of interest
VC: Validation Cohort
6MWT: Six-Minute Walk Test

