## Supplementary Appendix for "Derivation and validation of a non-invasive optoacoustic imaging biomarker for patients with intermittent claudication"

This supplementary appendix has been provided to give readers additional information about the work.

### Supplementary Material and Methods

##### Survey of study relevant medical history

In order to collect relevant medical information from the study participants, the electronic patient files and the collected information during anamnesis were used. In addition to general information, such as date of birth, sex, weight, height and skin type according to Fitzpatrick , vascular risk factors (smoking, arterial hypertension, dyslipidemia, diabetes mellitus, obesity with BMI>30kg/m^2^, positive family history) were identified. Previous performed vascular surgeries (interventional/open surgery/both), as well as important previous diseases, such as coronary artery disease, heart failure, atrial fibrillation, myocardial infarction, preterminal and terminal chronic kidney disease (with indication of the creatinine value), carotid stenosis and stroke were also determined. Furthermore, the use of relevant medications (antihypertensives, lipid-lowering agents, antidiabetics, acetylsalicylic acid, clopidogrel/ticagrelor, heparin, oral anticoagulation, coumarin, naftidroforyl, cilostazol, and prostanoids) was queried. Finally, the participants were asked about their subjective assessment of how far they could walk until the first pain occurres (relative walking distance), or until they have to stop for the first time due to pain (absolute walking distance).

##### Heel raise exercise

The heel raise exercise is designed to achieve maximal load and exhaustion of the calf muscle. For this purpose, the subject is asked to change from the normal stance to the toe-ball stance for at least 30 seconds at 1-second intervals. If there is no pain and no fatigue after 30 seconds in calf muscle, the subject should continue to alternate between normal stance and toe-ball stance until either event occurs. Figure 5 shows a schematic illustration of the heel raise exercise.

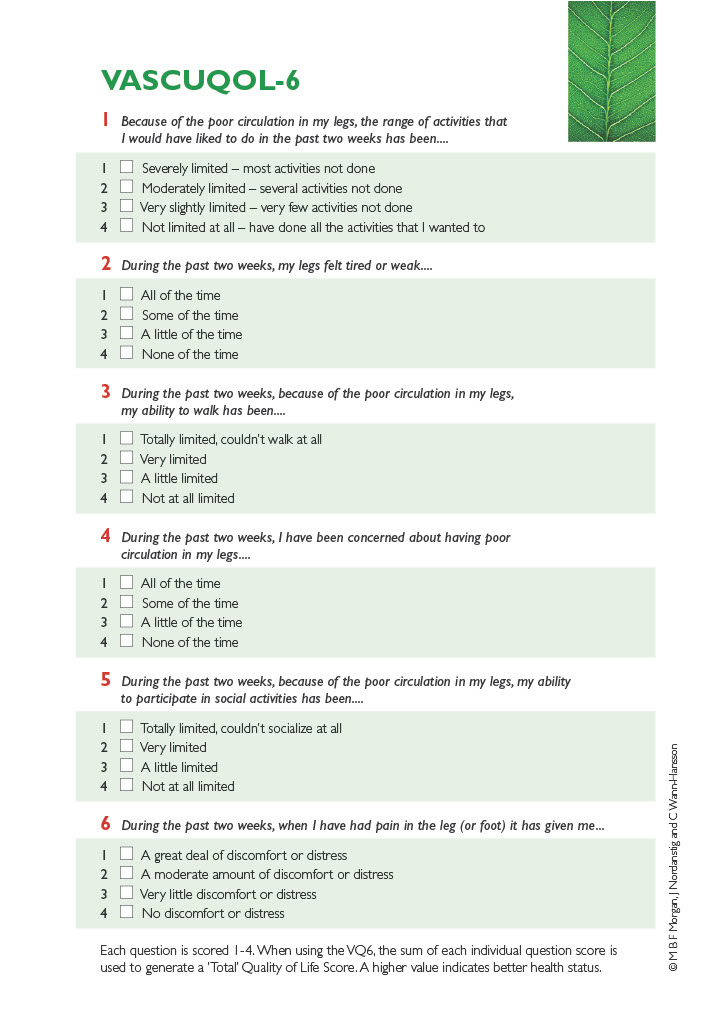

### Supplementary Figures

Figure S1 – VASCUQOL-6 questionnaire [1]

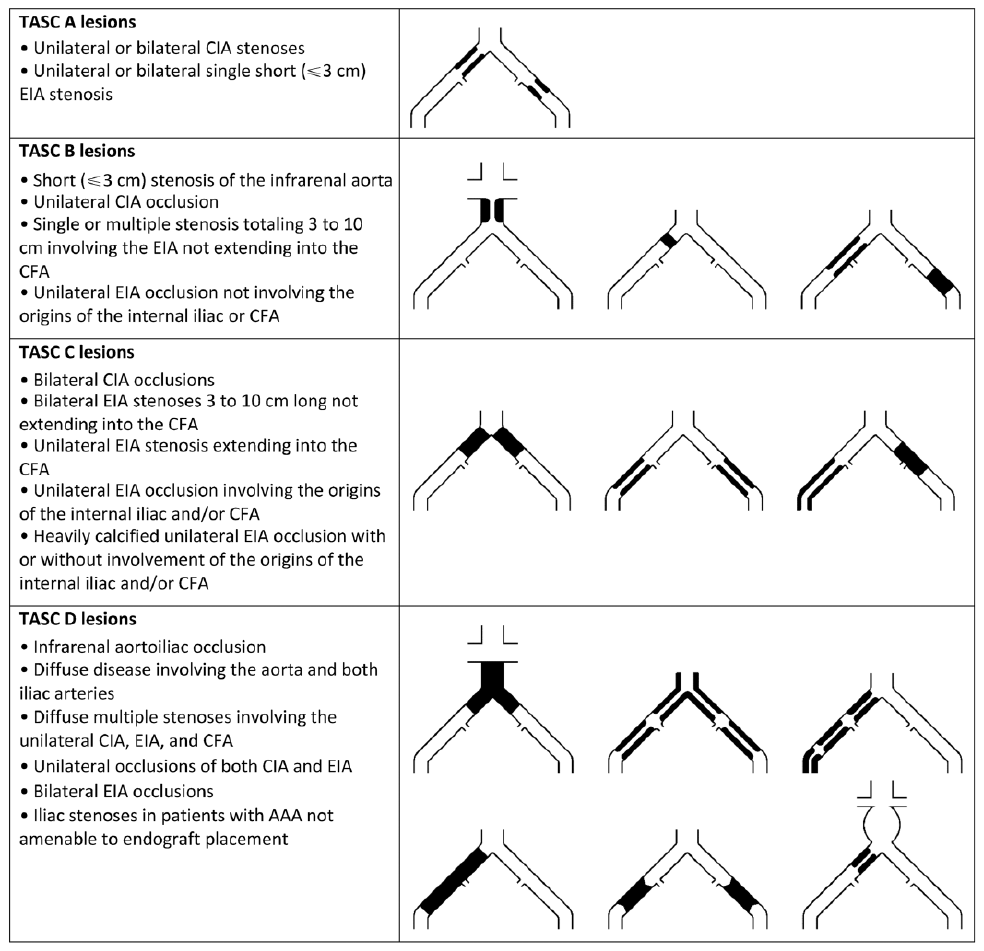

Figure S2 – TASC II classification: Aorto-iliac segment (AI) [2] (AAA, abdominal aortic aneurysm CFA, common femoral artery; CIA, common iliac artery; EIA, external iliac artery)

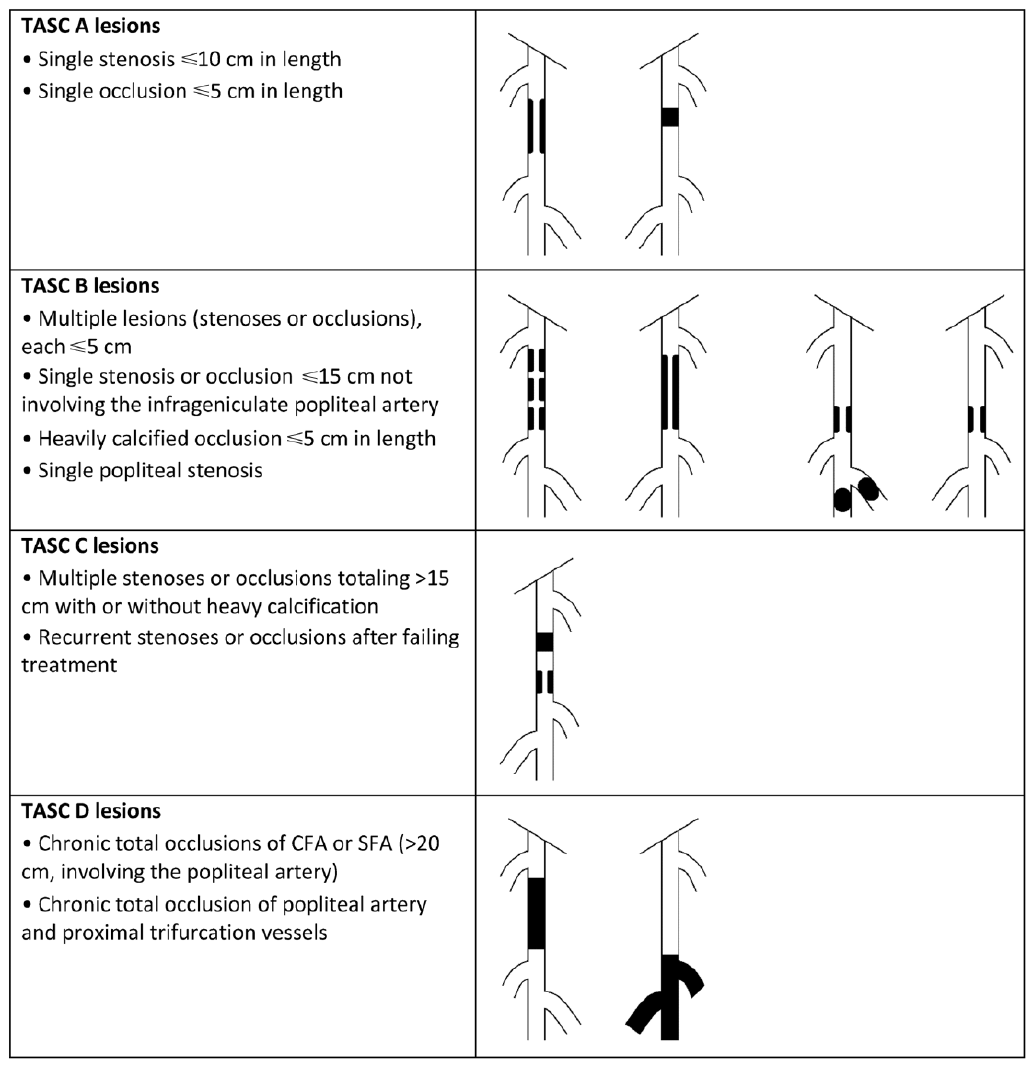

Figure S3 – TASC II classification: Femoral popliteal segment (FP) [2] (CFA, common femoral artery; SFA, superficial femoral artery)
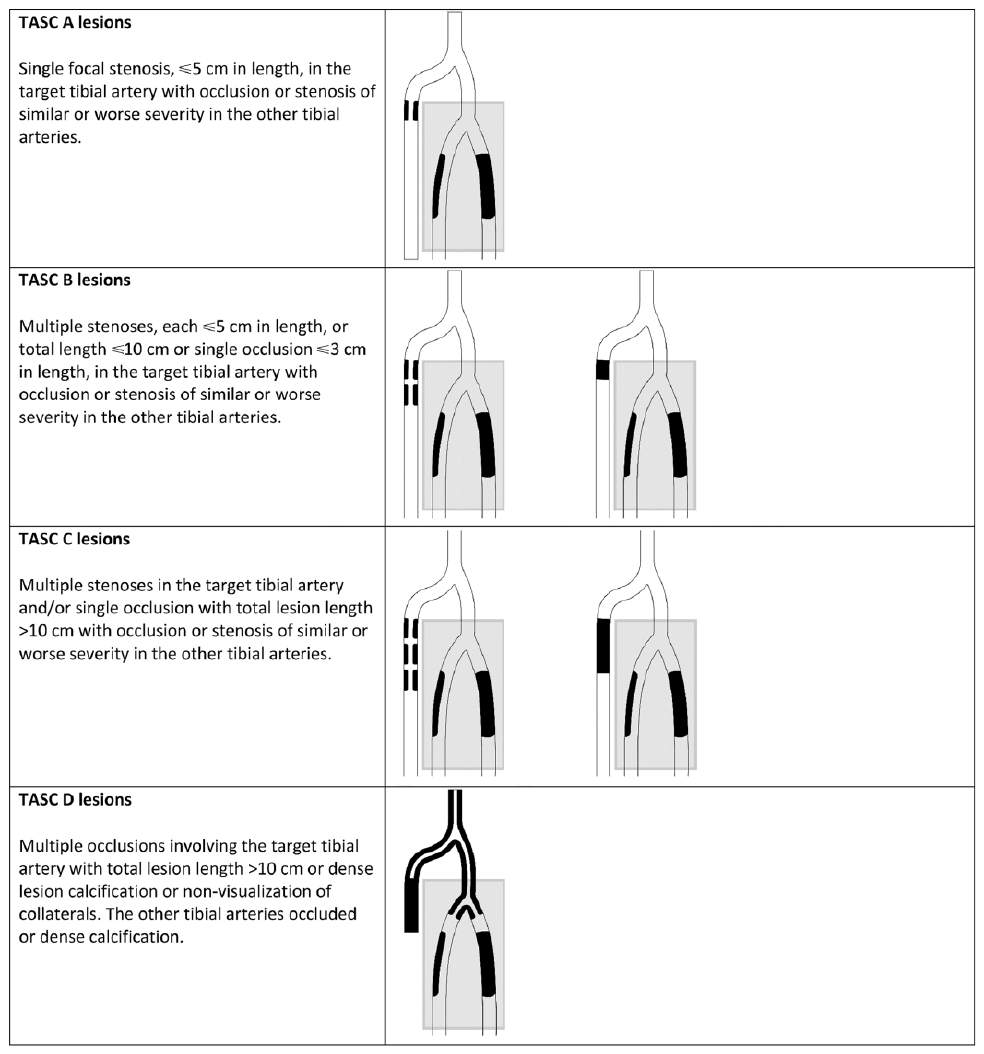

Figure S4 – TASC II classification: Infrapopliteal segment (IF) [3, 4]

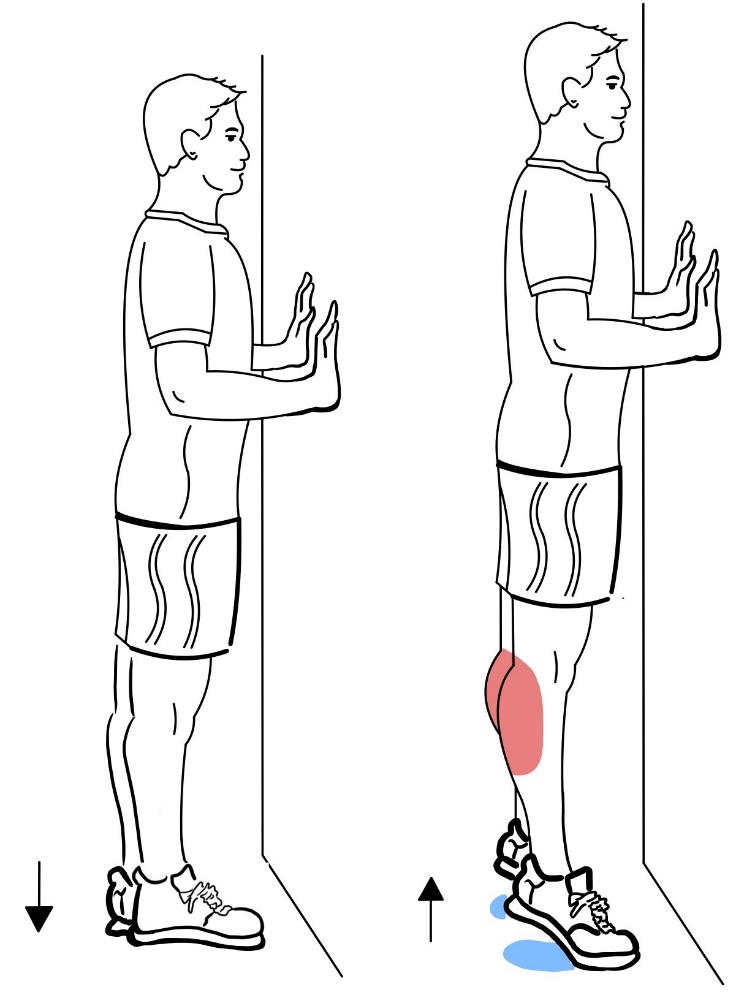

#### Figure S5 – Schematic illustration of the heel raise exercise

### Supplementary Tables

|  | aTASC 1 | aTASC 2 | aTASC 3 |
| --- | --- | --- | --- |
| Interpretation | angiographically healthy | good collateralization capability | poor collateralization capability |
| Findings according to TASC II classification | 1. no findings in the AI area  2. no findings in FP area  3. no findings in IP area | 1. no or A/B lesion in AI area  2. no or A/B/C/D lesion in FP area  3. any lesion in IP area  4. at least A/B lesions in 1. or 2. | 1. C/D lesion in AI area  2. no or A/B/C/D lesion in FP area  3. any lesion in IP area |

#### Table S1 – aTASC classification based on the TASC II classification

(TASC II, Trans-Atlantic Inter-Society Consensus Document on Management of Peripheral Arterial Disease classification II; aTASC, aggregated TASC II classification; AI, Aorto-iliac segment; FP, Femoral popliteal segment; IP, Infrapopliteal segment)

| Characteristic | Total cohort  (n = 102) | HV (n = 52) | | p-value | IC (n = 50) | | p-value |
| --- | --- | --- | --- | --- | --- | --- | --- |
|  |  | DC (n = 27) | VC (n = 25) |  | DC (n = 24) | VC (n = 26) |  |
| Age, yrs. | 63.83 ± 8.02 | 60.27 ± 6.66 | 59.64 ± 5.34 | .712 | 68.21 ± 7.91 | 67.49 ± 8.07 | .752 |
| Sex, n (%)  Female | 46 (45.1) | 18 (66.7) | 12 (48.0) | .173 | 5 (20.8) | 11 (42.3) | .104 |
| Risk factors, n (%)  Smoking  Arterial hypertension  Dyslipidemia  Diabetes mellitus  Obesity (BMI>30kg/m^2^)  Positive family history | 55 (53.9)  56 (54.9)  47 (46.1)  16 (15.7)  12 (11.8)  45 (44.1) | 2 (7.4)  6 (22.2)  11 (40.7)  0 (0.0)  1 (3.7)  10 (37.0) | 6 (24.0)  5 (20.0)  6 (24.0)  0 (0.0)  0 (0.0)  13 (52.0) | .098  .845  .199  N/A  .331  .278 | 24 (100.0)  21 (87.5)  14 (58.3)  7 (29.2)  4 (16.7)  11 (45.8) | 23 (88.5)  24 (92.3)  16 (61.5)  9 (34.6)  7 (26.9)  11 (42.3) | .086  .571  .817  .680  .382  .802 |
| Relevant diseases, n (%)  Coronary artery disease  Carotid stenosis  History of myocardial infarction  Stroke | 17 (16.7)  21 (20.6)  8 (7.8)  8 (7.8) | 0 (0.0)  0 (0.0)  0 (0.0)  0 (0.0) | 0 (0.0)  2 (8.0)  0 (0.0)  1 (4.0) | N/A  .134  N/A  .294 | 10 (41.7)  9 (37.5)  5 (20.8)  5 (20.8) | 7 (26.9)  10 (38.5)  3 (11.5)  2 (7.7) | .272  .944  .370  .181 |
| Current medication, n (%)  Antihypertensive  Lipid-lowering agent  Antidiabetic | 54 (52.9)  36 (35.3)  14 (13.7) | 6 (22.2)  2 (7.4)  0 (0.0) | 5 (20.0)  2 (8.0)  0 (0.0) | .845  .936  N/A | 18 (75.0)  15 (62.5)  5 (20.8) | 25 (96.2)  17 (65.4)  9 (34.6) | .031  .832  .278 |
| Previous revascularization procedure, n (%) | 22 (21.6) | 0 (0.0) | 0 (0.0) | N/A | 8.8 (33.3) | 14 (53.8) | .144 |
| Ankle-brachial-index  Before the exercise  After the exercise | 0.84 ± 0.32  0.72 ± 0.42 | 1.12 ± 0.06  1.07 ± 0.10 | 1.11 ± 0.06  1.10 ± 0.10 | 0.734  0.308 | 0.57 ± 0.26  0.35 ± 0.27 | 0.54 ± 0.17  0.31 ± 0.16 | 0.575  0.522 |
| VASCUQOL-6 score | 18.65 ± 6.20 | 24.00 ± 0.00 | 24.00 ± 0.00 | N/A | 13.42 ± 2.95 | 12.77 ± 5.07 | .581 |
| Walking distance in 6MWT [m]  Relative  Absolute  Total | 115 ± 71  207 ± 88  463 ± 142 | N/A  N/A  583 ± 34 | N/A  N/A  578 ± 52 | N/A  N/A  .649 | 109 ± 55  200 ± 99  342 ± 110 | 120 ± 84  213 ± 80  339 ± 88 | .594  .689  .914 |
| PAD stage according to Fontaine, n (%)  IIa  IIb | 24 (23.5)  26 (25.5) | 0 (0.0)  0 (0.0) | 0 (0.0)  0 (0.0) | N/A  N/A | 15 (62.5)  9 (37.5) | 9 (34.9)  17 (65.4) | .088  .088 |
| PAD stage according to aTASC, n (%)  aTASC 1  aTASC 2  aTASC 3 | 53 (52.0)  41 (40.2)  8 (7.8) | 27 (100.0)  0 (0.0)  0 (0.0) | 25 (100.0)  0 (0.0)  0 (0.0) | N/A  N/A  N/A | 1 (4.2)  18 (75.0)  5 (20.8) | 0 (0.0)  23 (88.5)  3 (11.5) | .480  .281  .305 |

#### Table S2 – Demographic and Clinical Characteristics

Categorical data represented by absolute and relative frequencies, continuous data by mean ± standard derivation. Statistical comparison of the DC and VC using the Chi-squared test for categorical data and the t-test for continuous data. There is no significant difference between the two groups in the data considered (p-value <0.05). (HV, healthy volunteers; IC, patients with peripheral arterial disease with in Fontaine stage IIa/IIb or Rutherford category 1 to 3; DC, derivation cohort; VC, validation cohort; VASCUQOL-6, Vascular Quality of Life Questionnaire-6; 6MWT, Six-Minute-Walk-Test; PAD, peripheral arterial disease; aTASC (aggregated TASC II classification; Type 1: HV or IC with no signs of stenosis or occlusion in angiography; Type 2: signs of stenosis or occlusion in the femoropopliteal and/or infrapopliteal area or TASC-II-level A or B in the aortoiliac area; Type 3: TASC-II-level C or D in the aortoiliac area); N/A, not applicable)

|  | DC | | | | | VC | |
| --- | --- | --- | --- | --- | --- | --- | --- |
|  | AUC | Sensitivity | Specificity | Youden index | Cut-off | Sensitivity | Specificity |
| Pre |  |  |  |  |  |  |  |
| Hb | 0.647 (0.495-0.799) | 0.889 (0.719-0962) | 0.250 (0.120-0.449) | 0.14 | 0.211 | 0.154 (0.062-0.225) | 0.880 (0.700-0.958) |
| HbO_2_ | 0.582 (0.424-0.749 | 0.296 (0.159-0.485 | 0.917 (0.742-0.985) | 0.21 | 0.232 | 0.885 (0.710-0.960) | 0.400 (0.234-0.593) |
| msO_2_ | 0.849 (0.741-0.956) | 0.815 (0.633-0.918) | 0.790 (0.595-0.908) | 0.61 | 0.525 | 0.769 (0.580-0.890) | 0.600 (0.407-0.766) |
| Post |  |  |  |  |  |  |  |
| Hb | 0.866 (0.766-0.965) | 0.741 (0.553-0.868) | 0.917 (0.742-0.985) | 0.66 | 0.199 | 0.769 (0.580-0.890) | 0.720 (0.524-0.857) |
| HbO_2_ | 0.867 (0.764-0.971) | 0.667 (0.478-0.814) | 1.000 (0.862-1.000) | 0.67 | 0.239 | 1.000 (0.871- 1.000) | 0.720 (0.524- 0.857) |
| msO_2_ | 0.991 (0.971-1.000) | 1.000 (0.875-1.000) | 0.958 (0.798-0.998) | 0.96 | 0.523 | 0.962 (0.811-0.998) | 0.960 (0.805-0.998) |

#### Table S3 – Representation and validation of the diagnostic quality of the MSOT measurement

The three MSOT parameters deoxygenated hemoglobin (Hb), oxygenated hemoglobin (HbO_2_) and MSOT saturation (msO_2_) are observed and analyzed before (Pre) and after (Post) the heel raise exercise. The area under the curve (AUC), sensitivity, specificity, and the cut-off point (calculated by using the optimal Youden index) are determined in the derivation cohort (DC). The cut-off point is applied in the validation cohort (VC) to validate the sensitivity and specificity calculated in the DC.
